# Movement-related beta modulation in amyotrophic lateral sclerosis depends on muscle strength: A magnetoencephalography study

**DOI:** 10.1101/2022.09.28.22280359

**Authors:** Tobias Sevelsted Stærmose, Lau Møller Andersen, Sarang S. Dalal, Christopher J. Bailey, Jakob Udby Blicher

## Abstract

**Background:** Movement related cortical beta (13-30 Hz) modulation is fundamental in the preparation and execution of movement. This oscillatory modulation is altered in amyotrophic lateral sclerosis (ALS) during active movement, with reports of both decreased and increased beta band power. These beta band changes have never been examined in a proprioceptive paradigm in ALS.

**Methods:** Using magnetoencephalography (MEG) we examined 11 ALS patients and 12 healthy participants. We recorded beta band activity during a session of active movement of the dominant hand index finger, using a visual cue. We also recorded activity during a passive movement of the same finger using a MEG compatible pneumatically activated device. All ALS patients underwent a clinical examination including an estimation of the muscle strength of the arm used for the experiment.

**Results:** Using an analysis of variance (ANOVA), we find that movement related beta band power is modified by ALS and the amplitude of beta power is decreased, both for the active and passive movements. We also find that the beta band power modulation depends on the muscle strength of the arm used, with movement related power amplitude being decrease in patients with arm weakness. This was observed for both active and passive movement.

**Conclusion:** ALS patients show decreased movement related beta band amplitude compared to the healthy control group. The decrease seems to depend on disease severity. These results show that ALS affects the motor outputs and sensory inputs of the sensorimotor cortex and that the modulation differs depending on disease severity. Severity dependent modulation of beta power could be related to disturbance in excitatory/inhibitory intracortical circuitry.

## Introduction

Amyotrophic lateral sclerosis (ALS) is a fatal progressive motor neuron disease that affects both cortical and spinal motor neurons. The pathophysiology and origin of ALS are still not fully understood (1). The diagnostic delay from first symptom to the diagnosis is long with few useful biomarkers. This usually means patients are first diagnosed at very varying stages of the disease (2).

ALS was previously regarded as a pure motor neuron disease (MND), now we know that multiple other areas and functions of the brain is involved, including prefrontal areas leading to symptoms of frontotemporal dementia (FTD) in up to 50% of ALS patients(3). Very little is known about the affection of the sensory function in ALS and the integration of sensory inputs to the sensorimotor areas of the brain in ALS is largely unexplored.

In this study we investigate the neural oscillations occurring in the beta band (13-30 Hz) of the sensorimotor cortex in both active and passive movement tasks, using magnetoencephalography (MEG). This enables us to investigate whether ALS impacts only the movement output or also leads to changes in processing of input to the sensorimotor cortex.

Both electroencephalography (EEG) and MEG allow for investigation of beta band oscillations non-invasively. The results from prior EEG and MEG studies investigating motor tasks in ALS are inconclusive on pathognomonic changes in the movement related beta band as highlighted by a recent review (4). In healthy subjects, motor activity, both actual movement and imaginary movement, lead to modulation of cortical oscillations in the *μ* (8–12 Hz) and *β* (13–30 Hz) bands in a well described manner with an event-related desynchronization (ERD) before the movement followed by event-related synchronization (ERS) after the movement has been finished (5, 6). The movement-related beta band has been examined in ALS compared to healthy controls (HC) by a handful of previous studies; most prior EEG results indicate reduced event-related changes in beta power in patients suffering from ALS (7-9). To our knowledge the only prior motor task studies in ALS using MEG are by Proudfoot et al. (10, 11), who show increased beta-ERD and delayed beta-ERS compared to a group of healthy controls.

Using MEG has some distinct advantages. It is a non-invasive and comfortable method for measuring cortical oscillations with high temporal precision (< 1 ms) and good spatial resolution (< 1 cm) (12)(13)To our knowledge, no prior studies have investigated the afferent features of the motor system in a passive movement task in ALS. The response to a passive finger movement has been shown to mainly contain proprioceptive information (14). Proprioception is highly important for the execution of motor function, encoding the position of the limbs of the body in its movement range. Passive movement activates sensory areas of the brain (15) and modulates 20 Hz beta band activity in motor areas (16).

We hypothesized that patients suffering from ALS will have altered cortical activity in the beta band during active movement compared to healthy subjects. Moreover, we hypothesized that this difference will be specific to active movement, whereas passive movement will induce similar changes in beta power in patients and healthy subjects. Finally, we hypothesized that the disease severity will interact with the cortical activity.

## Materials and methods

### Participants

ALS patients were recruited from the ALS clinic at the Neurology Department at Aarhus University Hospital, Denmark. All were diagnosed according to the latest revision of the El Escorial Criteria (17, 18). The healthy age- and gender-matched controls were excluded if they had a previous history of neurological or psychological disease or if they had limited movement abilities for any reason in their arms. All participants were given written and oral information before providing written informed consent. The study was approved by the scientific ethic committee of Region Midtjylland, Denmark (ID number: *65059*).

Thirteen healthy controls (HC) were recruited through an ad in the local newspaper “Favrskov Poster”, or from spouses and friends being informed of the study due to patient participation (no bloodline relatives). One control was excluded due to a later discovery of a history of multiple minor strokes leaving 12 subjects in the group for the analysis.

### Patient – Clinical examination

Disease severity was evaluated by a neurological examination as well as a rating of function using the revised ALS Functional Rating Score (19) (ALSFRS-R, range 0–48, lower scores reflecting greater disability). Muscles strength testing of 36 muscles groups were evaluated using the Medical Research Council scale (20) (MRC, 0-5). A subset of 11 muscles in the right upper extremity (RUE) was combined in a single score (RUE, includes dominant side (right side for all participants); *Shoulder abduction, shoulder adduction, elbow flexion, elbow extension, wrist flexion, wrist extension, finger flexion, finger extension, first dorsal interosseous, abductor digiti minimi and abductor pollicis brevis*). Upper motor neuron burden was evaluated using the Penn Upper Motor Neuron Score(21) (PUMNS, 0-32 points, higher scores – greater UMN burden).

The clinical and functional data were collected immediately after the MEG session or on a separate day within 2 weeks. All assessments were done by the same medical doctor (Author TSS), under supervision of an ALS expert neurologist (Author JB).

To investigate the effect of muscular strength on the beta band, patients were stratified based on the MRC Muscle Strength Scale score of their right upper extremity (RUE). Patients were classified as “minimally affected” (MA) if the total sum of MRC scores for the 11 RUE muscles was higher than 50 (maximum score 55), and “affected” (AF) if the total score was below 50. This resulted in two sub-groups:

### Task design

In order to get reliable ERD/ERS responses from both an active and passive motor task with a minimal number of other responses/tasks, a simple visual paradigm was designed: *Rest (including ready) (5-6 s), preparation (Yellow circle)= (2 s), action/move cue (green circle)(1 s), and back to rest*, (see Figure 1). The visual stimuli were back projected using a PROPixx projector (VPixx Technologies, Saint-Bruno, Canada) onto a screen approximately 1 m in front of the subject. The recordings were conducted in a dimly lit 3-layer magnetically shielded room (Vacuumschmelze GmbH & Co. KG, Hanau, Germany).

**Figure 1.**
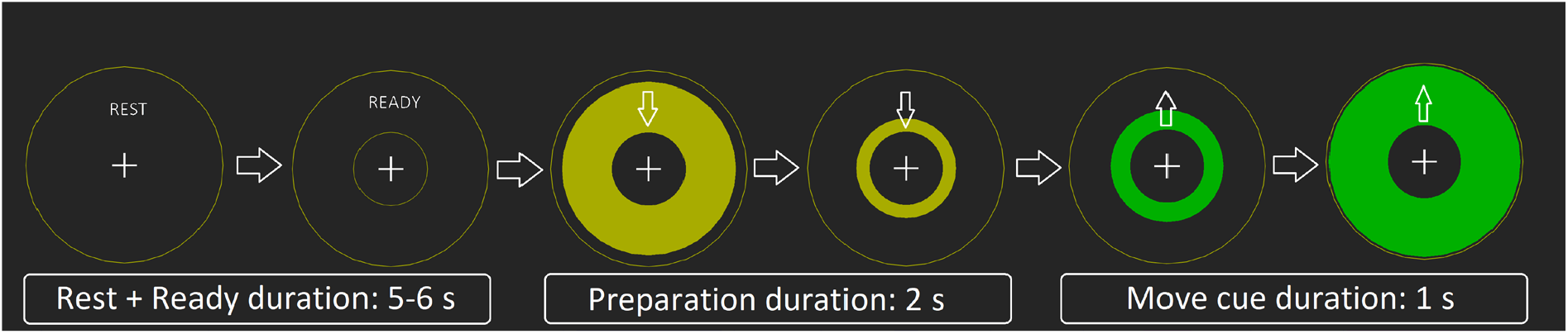
*The visual paradigm: From left to right; Subject focuses on the central cross while resting, “ready” indicates that soon the move task will commence. The yellow circle appears and moves in toward the central circle (2s), subject attempt to time their movement to when the yellow circle reaches the inner circle*. The green circle moves outwards (1s) indicating that the task is done and will return to rest, and then repeat.

In the active condition subjects were instructed to push a button against very light resistant foam (Figure 2a), using their dominant hand (all subjects right-handed) index finger following the visual cue. After extensive pilot testing we found that the 100 total movements were performed optimally with multiple breaks leading to a paradigm of 5×20 presses with 4×30 s breaks. All subjects were instructed to time the press of the button to the intersection of the yellow moving circle and the inner stationary circle. They were then told to press the button in a firm and decisive manner, “like a firm hand shake”, if the patient had little or no finger movement, they were still instructed the same even if the attempt to move produced no actual movement.

**Figure 2.**
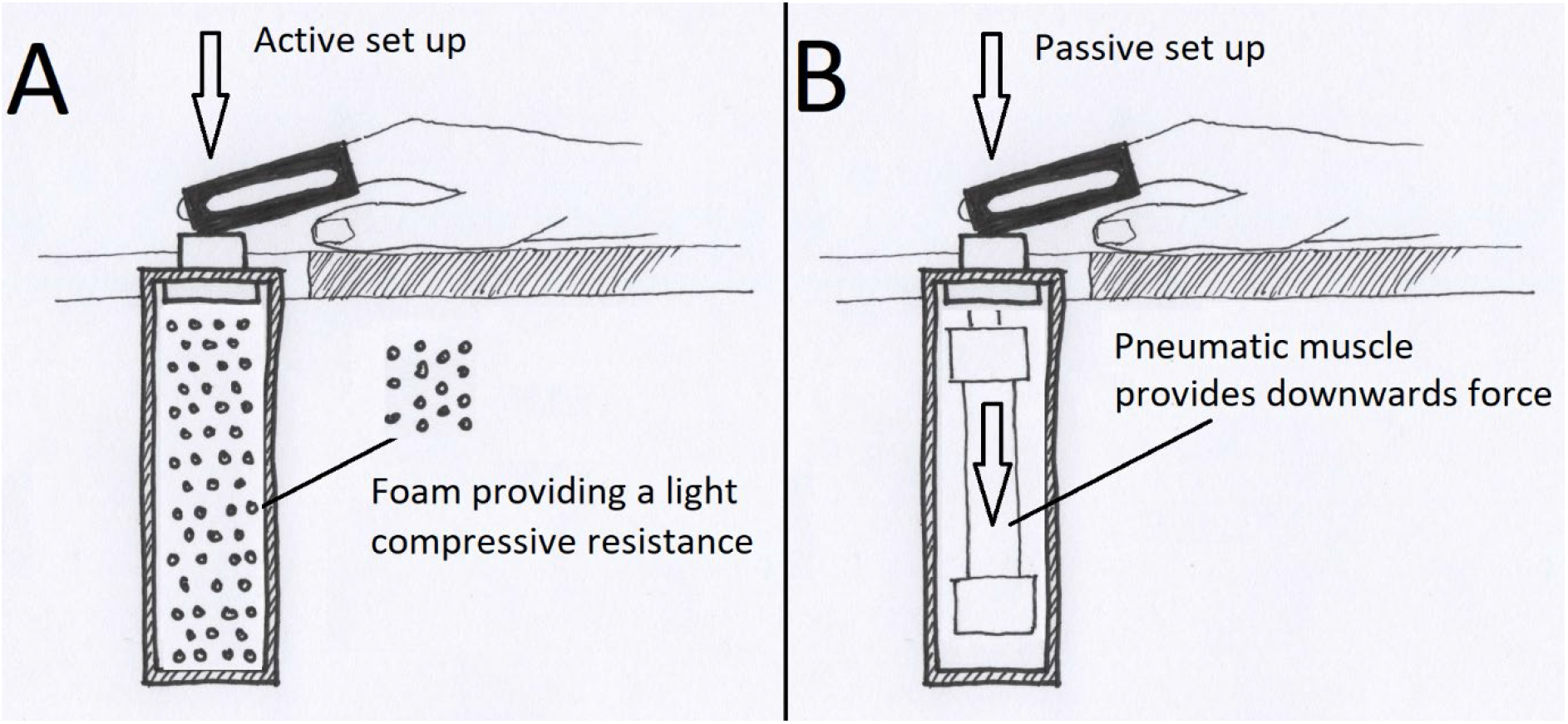
(A) Diagram of the active task. (B) Diagram of the passive task, the pneumatic controls connecting and actuating the muscle to the outside of the magnetically shielded room are not drawn. (Artist: Louise Sevelsted Stærmose)

The passive condition had an identical setup and visual cue. A pneumatic muscle (Fluidic Muscle DMSP, Festo, Germany) pulled the subject’s index finger to mimic the movement of the active condition (5×20 pulls, 4×30s breaks) (Figure 2b). Subjects were instructed to keep attention on the visual paradigm, but not move their finger.

The passive setup was based on the work of Piitulainen et al. (22), in our setup the subject had a relaxed position with their arms resting on a table in the standard position of the MEG compatible chair (Megin Oy, Finland). The pneumatic controls were provided by (Pnumatc Artificial Muscle Stimulator, Aalto NeuroImaging, Aalto University, Espoo, Finland). The active/passive button was mounted slightly above the table surface to place a slight extension on the subject’s index finger used for the experiment (see Figure 2).

In both conditions, the subject’s finger was placed in a hard plastic sheet to hinder the movement of the distal joints, and thus keep the movement to one joint, the metacarpophalangeal (MCP) joint.

To keep the arm and hand position the same in both the active and passive sessions, the active (foam filled) device was replaced with the passive pneumatic device in a small brake in the scan session while the subjects were still seated in the scanner.

### MEG acquisition

All subjects were scanned using a 306 channel Triux MEG system (Megin Oy, Finland). Simultaneous bipolar recordings of electromyography (EMG) from flexor digitorum profundus (FDP) and extensor indicis (EI) as well as right eye vertical electro-oculogram (EOG), using disposable gel-filled electrodes (Neuroline 720; Ambu A/S, Denmark) were collected during the scan. Prior to the MEG recording the subject had their head shape digitizedusing the Polhemus FASTRAK (Vermont, USA) system and 4 head position indicator coils were placed on the head of the subject to allow the MEG system to track head movement. An accelerometer (ACC) (ADXL335, Analog Devices, Inc., US) was mounted at the fingertip of the right index finger to record actual movement. The total time in the scanner was around 50 minutes. The MEG data were recorded at a 1000 Hz sampling rate with online 0.1 Hz high-pass and 330 Hz low pass filters.

### Data and Statistical analysis

Correction for head movement and noise was performed by preprocessing the raw data through MaxFilter software (Version v2.2.15; Elekta Oy, Helsinki, Finland). The MaxFiltered data was then further processed using MNE-Python v0.23.4 (23) in Python v 3.9.5. Eighteen gradiometer pairs covering the bilateral main motor and sensory areas in a 3×3 array were selected for a total of 36 channels, 18 for each hemisphere. The data was then filtered with independent component analysis (ICA) to remove any remaining EOG and cardiac artifacts using automatic MNE-Python functions. For each of the 100 active trials, per subject, the beginning of movement was manually marked based on the accelerometer data; this was used as the individual trial initiation for the further analysis of the active data (Movement Marker (MM)). Automatic movement correlations were tested but rejected due to inconsistent results. The MaxFiltered and ICA cleaned data was then epoched using MNE-Python.

Time 0 (t0) was set by the manual movement markers (MM) for the active condition. The passive condition had Time 0 (t0) locked at the paradigm trigger for the activation of the pneumatic muscle). The full time window was set as -4 s to 5 s from their respective condition t0.

General channel amplitude was used to reject epochs with max values of 300 pT/m for the gradiometers. This resulted in few epochs being rejected, typically 0-3 per100 epochs. The epochs were then used to compute Time-Frequency Representations (TFR) from 3-120 Hz using discrete prolate spheroidal sequences (DPSS) tapers using MNE-Python.

Three time windows were selected based on the group average TFR power (see Figure 4 for example). One series for a Rest comparison (RE), one for desynchronization (ERD), and one for the rebound (ERS). Slightly different time windows were used to capture the peak ERD/ERS amplitude window for each condition; for the active condition (ERD=-1.2 s-0.3 s, ERS=1.0 s-2.5 s) and the passive condition (ERD=-1.0 s-0.5 s, ERS=2.5 s-3.8 s) in both conditions a RE window was set to (−3.0 s to -2.0 s).

**Figure 3.**
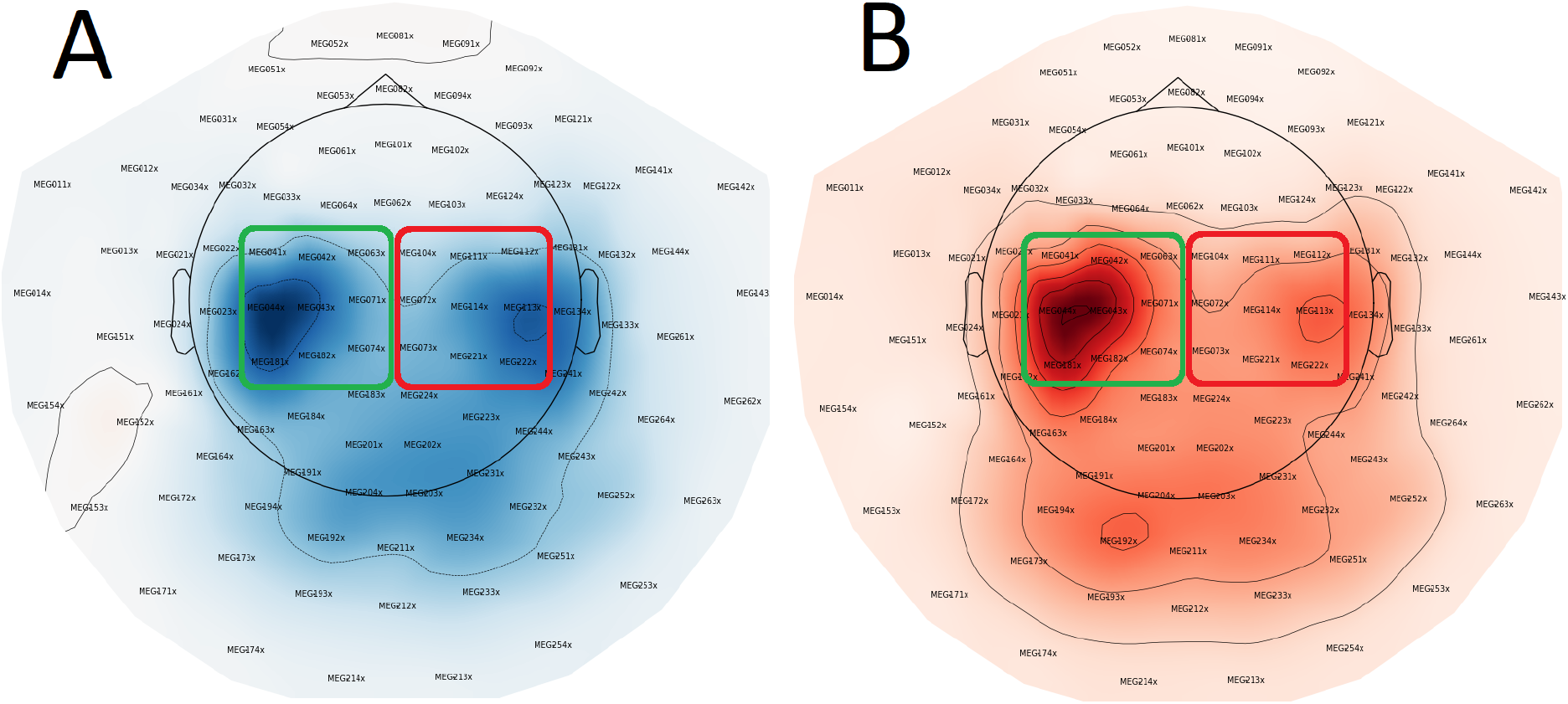
Visualization of selected sensors. Green box indicates contralateral sensors, while red indicates ipsilateral sensors. A: Controls, Active condition ERD, B: Controls Active condition ERS. Here data for the control group is shown, the ALS group shows very similar topographical activation.

**Figure 4.**
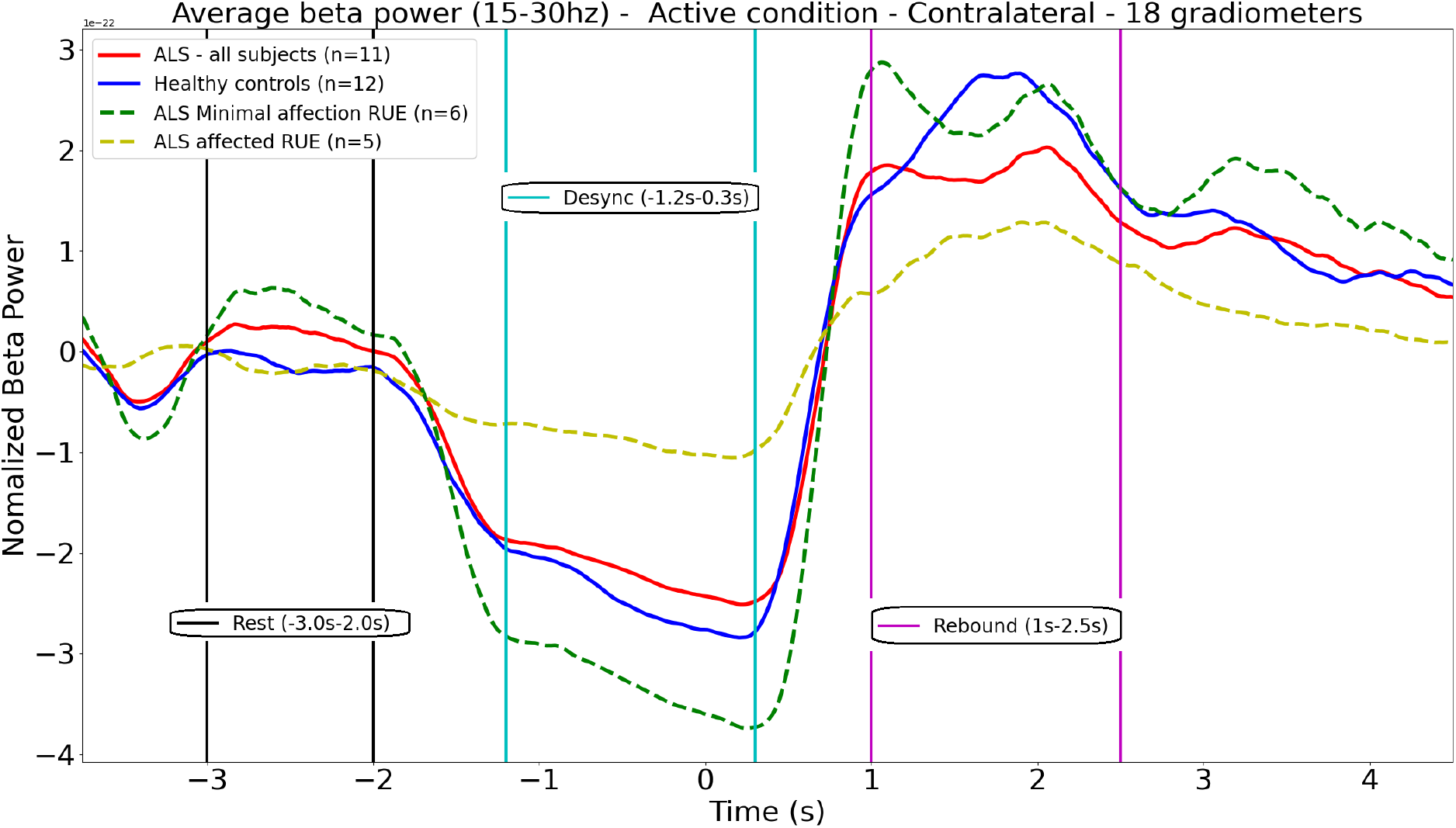
Normalized average beta power for the active condition, contralateral hemisphere. Vertical lines illustrate the time windows selected for the further analysis. t0 is the manual movement marker (MM).

An average power ratio (relative power (RP)) as the power in a given time window ratioed to the power of the full time window. This was then calculated for the individual TFRs for each subject, for the given conditions: active/passive, contralateral/ipsilateral, RE/ERD/ERS. This results in 12 average RP values per subject, as the combinations from investigating two hemispheres, condition active or passive, and baseline(rest), desync or rebound.

The power ratio calculated as average power for:

*RP = ((average power in “time window (eg ERD)”) / (total average power))*.

The total average power was calculated from the full TFR window subtracting 0.5 s in both ends to avoid artifacts from the filtering.

The final array was then imported to R (24) for further statistical analysis. In R we set up several linear models. The aim of the models are to examine the different variables and how much variance of the relative beta power is explained by the variables (as main effects or interactions effects). It is also examining what interactions, if any, are present between them and the relative beta power. If the relative power (RP) is influenced by the time window, the group of subject groups, the condition of the test or the hemispheres of the measurements we are able to test for these variables and any interaction between them using the analysis of variance (ANOVA) test. The alpha-level for significantly explaining variance in all models was *p* = 0.05.

***∼*** *= symbol dividing the dependent (left) and independent variables (right)*.

***\**** = signifies that the main effects and interactions of the independent variables are modeled.

The model for the group level (ALS / HC):

*(Relative beta power ∼ Time window (RE, ERD, ERS) * Group (ALS, HC) * Condition (Active, Passive) * Laterality (Contralateral, Ipsilateral))*.

The model for the strength stratification based on the type of motor function (RUE score) in the right arm (MA / AF / HC):

*(Relative beta power ∼ Time window (RE, ERD, ERS) * Type (MF, AF, HC) * Condition (Active, Passive) * Laterality (Contralateral, Ipsilateral))*.

To investigate if the effects are coming from the active or passive session we ran the analysis on a two-condition split. With the ANOVA models set for the group level as:

*(Relative beta power ∼ Time window (RE, ERD, ERS) * Type (ALS, HC) * Laterality (Contralateral, Ipsilateral))*.

And the RUE score stratified model as:

*(Relative beta power ∼ Time window (RE, ERD, ERS) * Type (MF, AF, HC) * Laterality (Contralateral, Ipsilateral))*.

In addition, we did correlations test using Pearson’s correlation coefficient for correlations of key areas in the paradigm and the clinical measurements of the patients group. We examined the correlation between clinical scores (ALSFRSr and PUMNS) and the contralateral beta ERD and ERS in both active and passive conditions.

## Results

When comparing HC with the ALS group (see table 3) we find a significant interaction between the group and the time windows; ALS significantly interacts with the beta-band power in rest, ERD and ERS compared to HC (*p* = 0.024, F = 3.788, degrees of freedom = 2). We find an interaction effect of the condition (active / passive) and the time windows on the beta-band power (*p* = 0.032, F = 3.505, degrees of freedom = 2). The laterality of the measurements also interacts with the time windows for the beta band power (*p* < 0.001, F = 18.081, degrees of freedom = 2).

**Table 1.** Participants. One ALS patient was carriers of the C9ORF72 mutation. ALS Functional Rating Score (ALSFRS-r). Penn Upper Motor Neuron Score (PUMNS). Medical Research Council scale (MRC) for muscle strength in the right upper extremity (RUE).

| Mean ± SD (Range) | ALS (n=11) | Controls(n=12) |
| --- | --- | --- |
| Age (Years) | 57.8 ± 16.3 (33-75) | 58.7 ± 15.0 (31-77) |
| Gender | 4F / 7M | 6F / 6M |
| Handedness | 11 Right | 12 Right |
| ALSFRS-r | 36.3 ± 7.7 (22-45)* | N/A |
| Time since diagnosis (months) | 12.5 ± 10.7 (1-41)* | N/A |
| PUMNS (32 points max, lower is better) | 6.5 ± 8.0 (0-28) | N/A |
| Right arm strength score MRC - RUE (11 muscles, 55 points max) | 45.5±11.4 (19-55) | N/A |
| Riluzole treatment active at the time of the study | 7 Yes / 3 No* | N/A |
| * One data set incomplete, data on 10/11 ALS subjects. |  |  |

**Table 2.** Stratifications based on right upper extremity MRC muscle strength scores.

|  |  |
| --- | --- |
| Minimally affected right arm | N=6 (average total MRC score = 54.5/55, range: 53-55) |
| Affected right arm | N=5 (average total MRC score = 34.6/55, range 19-42) |

**Table 3.**
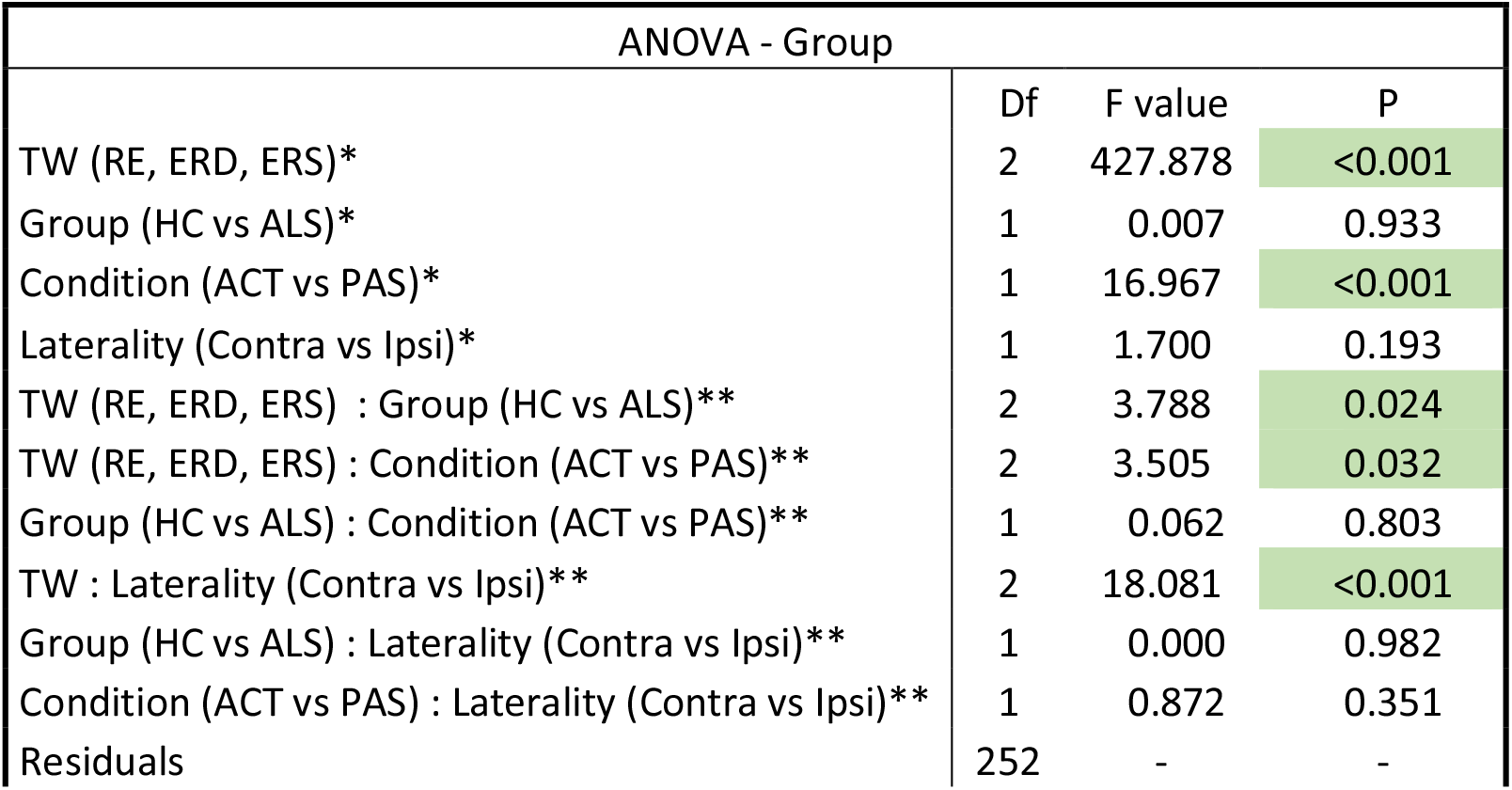
Results of the ANOVA test on the group-based model. 3- and 4-way interactions not shown, all p-values >0.652. * Main effects. ** Two-way interactions. Time window (TW) for; Rest (RE), event related desynchronization (ERD) and event related synchronization (ERS). Conditions; Active (ACT) and Passive (PAS). Group; Healthy controls (HC) and ALS patients (ALS). Laterality; the hemisphere of measurements, contralateral (Contra) and ipsilateral (Ipsi). Degrees of freedom(Df)

| ANOVA - Group |  |  |  |
| --- | --- | --- | --- |
|  | Df | F value | P |
| TW (RE, ERD, ERS)* | 2 | 427.878 | <0.001 |
| Group (HC vs ALS)* | 1 | 0.007 | 0.933 |
| Condition (ACT vs PAS)* | 1 | 16.967 | <0.001 |
| Laterality (Contra vs Ipsi)* | 1 | 1.700 | 0.193 |
| TW (RE, ERD, ERS) : Group (HC vs ALS)** | 2 | 3.788 | 0.024 |
| TW (RE, ERD, ERS) : Condition (ACT vs PAS)** | 2 | 3.505 | 0.032 |
| Group (HC vs ALS) : Condition (ACT vs PAS)** | 1 | 0.062 | 0.803 |
| TW : Laterality (Contra vs Ipsi)** | 2 | 18.081 | <0.001 |
| Group (HC vs ALS) : Laterality (Contra vs Ipsi)** | 1 | 0.000 | 0.982 |
| Condition (ACT vs PAS) : Laterality (Contra vs Ipsi)** | 1 | 0.872 | 0.351 |
| Residuals | 252 | - | - |

When stratifying on the right upper extremity strength score (RUE score) (Minimally affected / Affected / HC) (see Table 4), we find a significant interaction on the type of RUE (MA/AF) and the time windows with the beta-band power in rest, ERD and ERS compared to HC (*p* < 0.001, F = 7.119, degrees of freedom = 4). In this model we again find the interaction of condition and laterality with the time windows on the beta power, with (*p* = 0.026, F = 3.703, degrees of freedom = 2) and (*p* < 0.001, F = 19.103, degrees of freedom = 2) respectively.

**Table 4.** Results of the ANOVA test on the strength-stratification-based model. 3- and 4-way interactions not shown, all p-values >0.482. * Main effects. ** Two-way interactions. Time window (TW) for; Rest (RE), event related desynchronization (ERD) and event related synchronization (ERS). Conditions; Active (ACT) and Passive (PAS). Group; Healthy controls (HC) and ALS patients. Type; ALS in two groups based on their right arm strength score (RUE score), minimally affected patients (MA) and affected patients (AF). Laterality; the hemisphere of measurements, contralateral (Contra) and Ipsilateral (Ipsi). Degrees of freedom(Df)

| ANOVA – Strength stratification |  |  |  |
| --- | --- | --- | --- |
|  | Df | F Value | P |
| TW (RE, ERD, ERS)* | 2 | 452.056 | <0.001 |
| Type (HC vs MA vs FA)* | 2 | 0.270 | 0.763 |
| Condition (ACT vs PAS)* | 1 | 17.926 | <0.001 |
| Laterality (Contra vs Ipsi)* | 1 | 1.796 | 0.181 |
| TW (RE, ERD, ERS) : Type (HC vs MA vs AF)** | 4 | 7.119 | <0.001 |
| TW (RE, ERD, ERS) : Condition (ACT vs PAS)** | 2 | 3.703 | 0.026 |
| Type (HC vs MA vs AF) : Condition (ACT vs PAS)** | 2 | 0.731 | 0.483 |
| TW : Laterality (Contra vs Ipsi)** | 2 | 19.103 | <0.001 |
| Type (HC vs MA vs AF) : Laterality (Contra vs Ipsi)** | 2 | 0.028 | 0.972 |
| Condition (ACT vs PAS) : Laterality (Contra vs Ipsi)** | 1 | 0.922 | 0.338 |
| Residuals | 240 | - | - |

Main effects from both models shown in Tables 3 and 4. A visual representation of the data showing the interaction between the ERD, ERS and the group of the subjects, see Figure 5.

**Figure 5.**
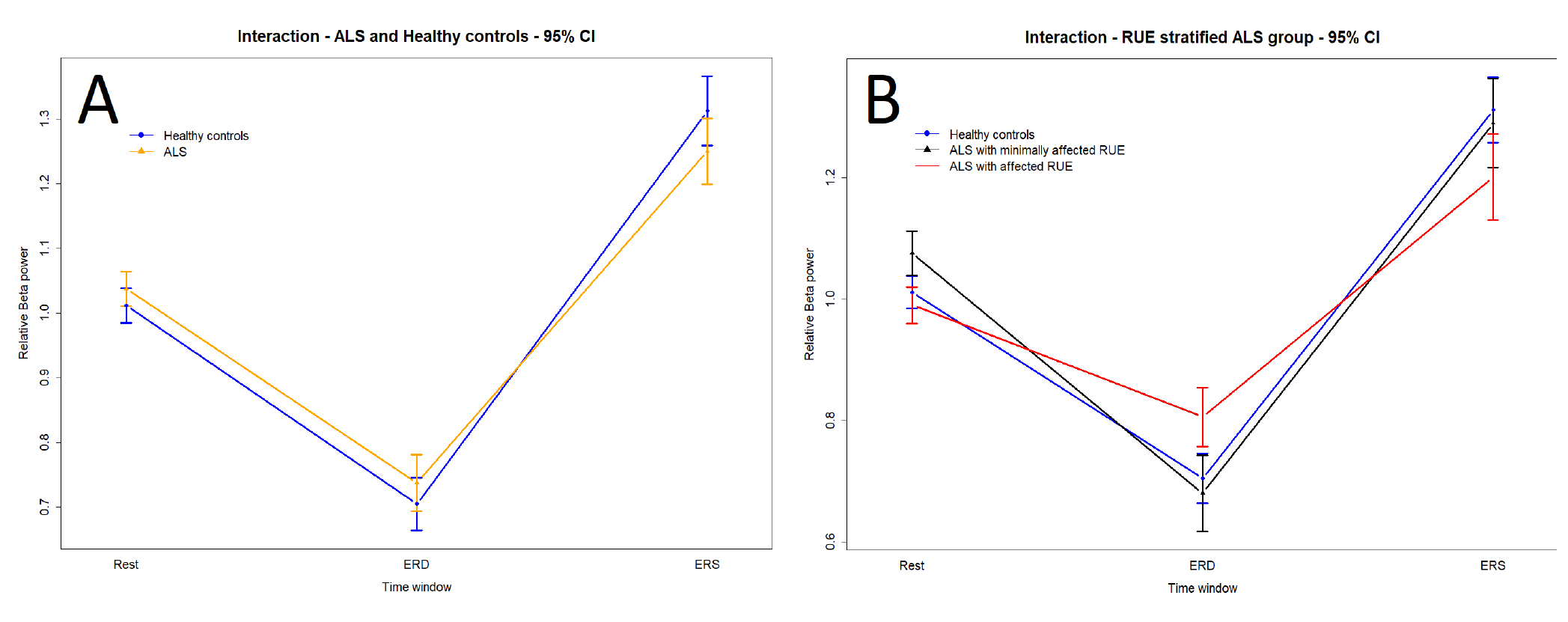
(A) The average Beta band power in the model for ALS-HC, over the 3 time windows. (B) The average Beta band power with patients stratified based on muscle strength (RUE stratified group). Error bars depict the 95% Confidence interval.

### Analysis for individual conditions

When analyzing each condition (active or passive) alone, we find a very similar pattern between active and passive movement, with a significant main effect of Time (RE, ERD, ERS) and a significant interaction between Time and Laterality. When comparing healthy subjects and patients we find no significant difference (Main effect of Group), and no significant interaction between Time and Group, that is no difference between when comparing healthy subjects to the combined patient group (indicating similar beta band power modulation between patients and healthy subjects). However, for both active and passive conditions we found a significant interaction between Time (RE, ERD, ERS) and Type (patients stratified based on muscle strength) indicating a difference between the groups when we account for loss of muscle strength in the patient group, and that this difference is present for both active and passive movement (see Table 5 and Figure 6).

**Table 5:** ANOVA results from separate active and passive conditions. No additional main effects or interactions were significant. * Main effects. ** Two-way interactions. Time window (TW) for; Rest (RE), event related desynchronization (ERD) and event related synchronization (ERS). Group; Healthy controls (HC) and ALS patients. Type; ALS patients in two groups based on their right arm strength score (RUE score), minimally affected patients (MA) and affected patients (AF). Laterality; the hemisphere of measurements, contralateral (Contra) and Ipsilateral (Ipsi). Degrees of freedom(Df).

| ANOVA - Group (ALS / HC) - Active condition |  |  |  |
| --- | --- | --- | --- |
|  | Df | F value | P |
| TW (RE, ERD, ERS)* | 2 | 231.127 | <0.001 |
| TW (RE, ERD, ERS):Group (HC vs ALS)** | 2 | 1.819 | 0.166 |
| TW (RE, ERD, ERS):Laterality (Contra vs Ipsi)** | 2 | 8.141 | <0.001 |
| Residuals | 126 |  |  |
| ANOVA - Strength stratification (HC / MA / AF) - Active condition |  |  |  |
|  | Df | F value | P |
| TW (RE, ERD, ERS)* | 2 | 242.112 | <0.001 |
| TW (RE, ERD, ERS):Type (HC vs MA vs AF)** | 4 | 3.632 | 0.008 |
| TW (RE, ERD, ERS):Laterality (Contra vs Ipsi)** | 2 | 8.528 | <0.001 |
| Residuals | 120 |  |  |
| ANOVA - Group (ALS / HC) - Passive condition |  |  |  |
|  | Df | F value | P |
| TW (RE, ERD, ERS)* | 2 | 198.982 | <0.001 |
| TW (RE, ERD, ERS):Group (HC vs ALS)** | 2 | 2.045 | 0.134 |
| TW (RE, ERD, ERS):Laterality (Contra vs Ipsi)** | 2 | 10.155 | <0.001 |
| Residuals | 126 |  |  |
| ANOVA - Strength stratification (HC / MA / AF) - Passive condition |  |  |  |
|  | Df | F value | P |
| TW (RE, ERD, ERS)* | 2 | 212.196 | <0.001 |
| TW (RE, ERD, ERS):Type (HC vs MA vs AF)** | 4 | 3.760 | 0.006 |
| TW (RE, ERD, ERS):Laterality (Contra vs Ipsi)** | 2 | 10.829 | <0.001 |
| Residuals | 120 |  |  |

**Figure 6.**
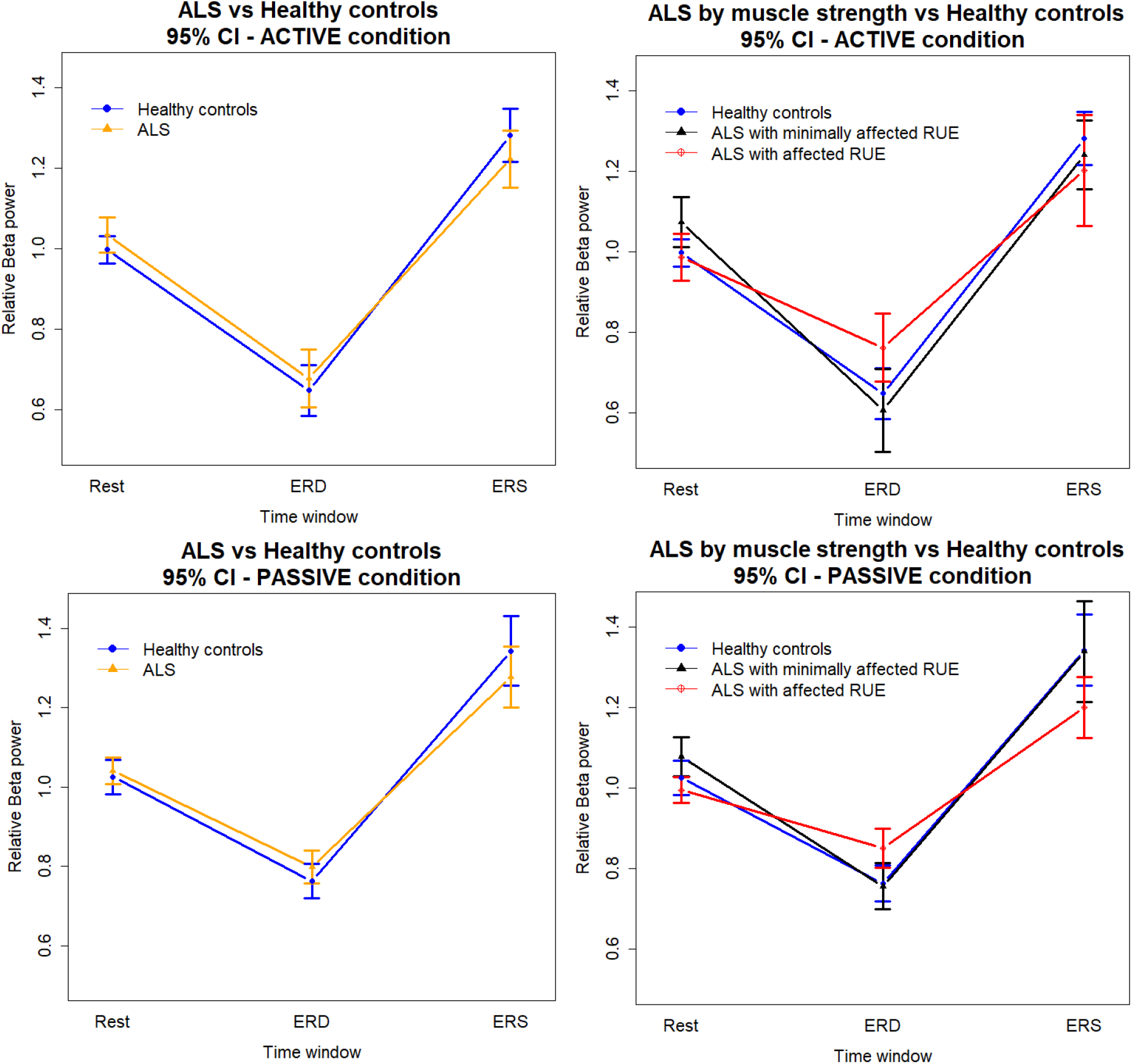
Interactions plot for the active and passive conditions, on both group and muscle strength of the right upper extremity score (RUE) stratified type of ALS patients. The interaction between groups are not significant, but a significant interaction is present for the patient group with affected arm strength (RUE).

### Correlation to clinical measures

The results from the correlation analysis are shown in Table 6 for the ALSFRSr results and Table 7 for the PUMN results. No significant correlations were found. See example plots in Figure 7.

**Table 2.** Pearson’s correlation coefficient. Conditions by time windows for event related desynchronization (ERD) and event related synchronization (ERS) and the ALSFRS-r scores. * See Figure 4A for example correlation plot.

| Correlation: ALSFRS-r - Contralateral Beta power |  |  |  |  |  |
| --- | --- | --- | --- | --- | --- |
| Condition | Time | Correlation | 95% CI Lower | 95% CI Upper | P-value |
| Active | ERD | -0.235 | -0.753 | 0.463 | 0.514 |
| Active* | ERS | 0.490 | -0.202 | 0.856 | 0.150 |
| Passive | ERD | -0.337 | -0.797 | 0.372 | 0.342 |
| Passive | ERS | 0.430 | -0.273 | 0.834 | 0.215 |

**Table 3.** Pearson’s correlation coefficient. Conditions by time windows for event related desynchronization (ERD) and event related synchronization (ERS) and the ALSFRS-r scores. * See Figure 4B for example correlation plot.

| Correlation: PUMNS - Contralateral Beta power |  |  |  |  |  |
| --- | --- | --- | --- | --- | --- |
| Condition | Time | Correlation | 95% CI Lower | 95% CI Upper | P-value |
| Active | ERD | 0.017 | -0.589 | 0.611 | 0.960 |
| Active* | ERS | -0.567 | -0.871 | 0.050 | 0.069 |
| Passive | ERD | 0.158 | -0.488 | 0.692 | 0.642 |
| Passive | ERS | -0.305 | -0.765 | 0.361 | 0.362 |

**Figure 7.**
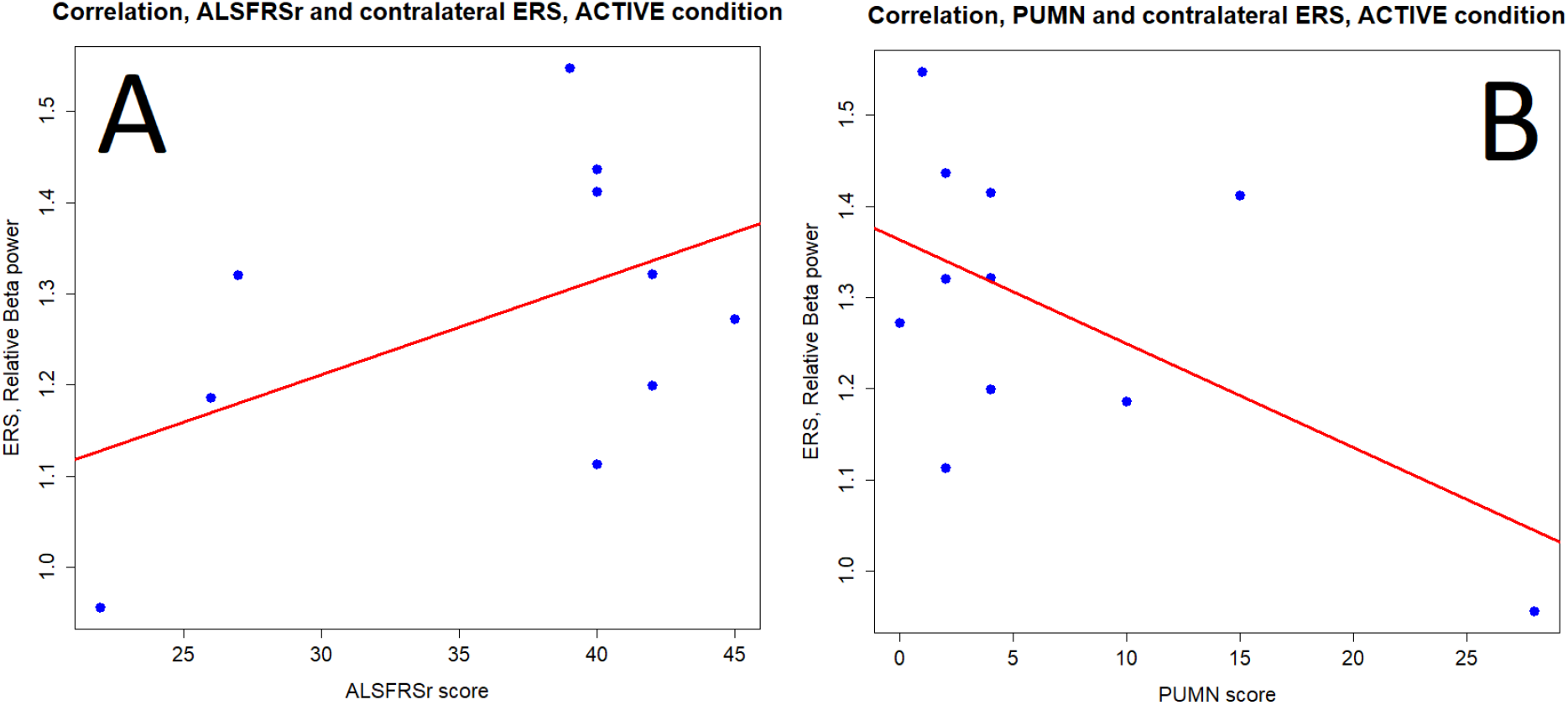
Examples of correlation of clinical scores and relative beta power. For both P>0.05. See Tables 6 and 7 for more.

## Discussion

Our results show that movement-related beta-band power is attenuated in ALS as opposed to healthy subjects.

When comparing the movement related beta band in ALS patients with that in healthy controls we find significant decreases in the amplitude of both beta ERD and ERS in a model containing both active and passive movement. This effect seems driven by the loss of muscle strength in the ALS patients.

Our results on beta power during active movement is overall similar to previous studies reporting; reduced beta-ERS (7), reduced ERD (8) and increased ERD and slowed ERS (10). Since these previous results have not been in full agreement, it is hard to conclude on pathognomonic ERD/ERS changes in ALS.

We had hypothesized that these changes in beta power compared to the healthy controls would only apply to the active movement output, not the input from a passive movement eliciting a proprioceptive response. To our surprise, beta power changes related to a passive movement differed between ALS and healthy subjects in a similar way as an active movement. For both active and passive movement, ERD and ERS movement related responses are less pronounced in the more severely affected ALS group. It would therefore seem that ALS is affecting the sensorimotor areas more widely. The cortical input from the passive condition is seemingly affected in the beta band in a similar way as in the active task that combines motor output and afferent feedback. The changes in both the active and passive tasks could suggest that change in the cortical excitability is a factor in our findings. Excitability can be modulated by change in the Gamma-aminobutyric acid (GABA) / glutamate balance in the brain (25). It is also known that the GABA concentration modulates the beta frequencies and power in the sensorimotor areas of the brain (26, 27). Furthermore it has been shown that the pre-execution part of a movement (ERD) and the reaffirmation of limb position after a movement (ERS) reacts differently to drugs modulating the GABA receptor (28) this would perhaps suggest that our finding are modulated by more than just GABA. A glutamate/GABA imbalance and hyperexcitability has been hypothesized as the cause of ALS. Several studies have investigated these neurotransmitters using magnetic resonance spectroscopy with some evidence for elevated glutamate and decreased GABA levels, suggesting a piece in the puzzle of neuronal excitability in ALS (29)

To further understand the interaction of ALS on the beta frequencies in our data we investigated the relationship between ERD/ERS changes and motor function in ALS and created a simple strength stratification of our ALS group based on the strength of the investigated limb. The results show that beta power for both ERD and ERS, changes depending on the severity of muscle deficits in the tested limb (right arm), likely driven by the progression of the disease. The affected group seemingly being the driver of the decrease in amplitude for both ERD and ERS compared to the less affected group as illustrated by the plots in Figure 7. With the MA group not showing the same changes to the beta band as the AF ALS group and not significantly deviating from the control group. Although Proudfoot et al. show excessive beta ERD (30) we do not find significant evidence for increased ERD in our groups.

The notion that ALS patients have changes in the cortical excitability as a function of disease progression have previously been suggested before from connectivity studies; a review by Agosta et al. (2018) reports on several studies that use resting state functional MRI to show both decreased functional connectivity (FC) of the sensorimotor networks as well as increased, or as a regional mix (31). Studies of resting state investigated with MEG/EEG have shown an increase in broad band FC(32, 33) and some showing a reduction in alpha (8-12 Hz) FC(34). This still leaves a complex picture of the changes in the excitability, neurotransmitters and oscillatory power caused by ALS. This phenomenon should be further investigated; if earlier disease stages have hints of increased excitability, and this could potentially be used for early prediction of ALS.

Our results also, as expected, show that beta band power depends on both condition of the task (active or passive) that is being performed (35) and differs between the two hemispheres (36-38).

Our results do not find a compelling correlation between the clinical measures of ALSFRS-r and PUMNS. Since the measures of beta power in this study are defined relative to a movement of the finger, it does not seem that either clinical scale captures the same state of ALS that relates to the beta ERD/ERS as the very specific nature of the RUE score we presented. With more subjects, it would possibly be more feasible to investigate potential correlation between beta power and ALSFRS-r/PUMNS.

One possible confounding factor driving the changes in ERD and ERS during the active task, could be the result of the effective force and speed that the subjects are able to generate, with the worse ALS patients being unable to produce the wanted force or speed on the active button press, or having to apply more effort to perform the task. The association between speed and ERD/ERS has been studied in healthy controls, with no changes in the ERD/ERS magnitude as a result of movement speed found by several studies (38, 39). However, one 2020 study by Zhang et al. found a modulation on the ERS amplitude depending on the speed of movement(40). The force of the movement has also been investigated with no changes in beta power (41), or the beta ERD but increased amplitude of the ERS (42). A 2019 study by Tatti et al. suggests that beta ERS and ERD is not modulated by movement features (force, speed, etc.). We cannot rule out that movement force, speed and subject effort could contribute to the changes that we find, but since they are present and similar in the passive condition, that should be mechanically similar for all subjects, we believe that it is more likely a feature of ALS, than a bias related to task performance.

The progressive nature of ALS is often explained by either a dying-forward (DF) or a dying-back (DB) hypothesis. The DF model suggests the involvement of the upper motor neurons (UMN) as the initial step in the disease. Whereas the DB model hypothesizes that the disease originates in the muscles or at the neuromuscular junction and subsequently lead to denervation and retrograde affection of the lower motor neurons (LMN) and then the UMN(1). Our results could potentially be evidence for the DB model as we do not find compelling evidence for cortical changes in the patient group with limited muscular deficits. If the beta power changes were present in the group that have not yet lost significant strength it would more likely be in favor of a DF model. The results from the MA group are not significant, but it could be speculated that it hints at an increase in activity in the brain before the muscles are severely affected, this would argue in favor of the DF model.

The number of patients in our study limits the ability to more precisely stratify patients according to muscle strength and how it interacts with the movement related beta power. More patients would allow for finer stratification and could have enabled a better understanding of whether or not the oscillatory changes are progressing linearly or not. Further longitudinal studies should be designed to elaborate on this change in beta power of the motor cortex driven by disease progression.

Exactly why the beta rhythms in the sensorimotor areas are influenced by ALS on both the output and input side is beyond this study. The overall excitability of the diseased brain could explain part of our findings; if further disease progression modulates and lowers excitability it could be a factor in the lower beta amplitude of the most affected ALS group.

Even with the limited number of patients our conclusions remain that ALS affects the movement-related beta band in both a motor execution task and during a passive movement.

We can also conclude that stratifying the patients by muscular strength is important as cortical changes in the beta band depend on muscle strength. Previous studies are somewhat inconclusive on pathognomonic ERD/ERS changes; this could in part be due to heterogeneous patient populations.

With our new suggestions for strength stratification, it would be possible to reanalyze results from previous studies if the required MRC data of patients is available. This would enable better understanding of the muscle strength and beta band without the need for new data to be collected. We also see this as piece in the puzzle of ALS progression and would be excited to see longitudinal MEG studies of ALS incorporate some form of MRS to directly follow the GABA and glutamate ratios together with beta band modulations, crucially making it possible to link them with the clinical performance of patients.

## Data Availability

All data produced in the present study are available upon reasonable request to the authors, as long as they can be considerd within the GDPR data protection framework.

## Funding

This study was supported by the Innovation Fund Denmark with a grant for the REMAP project (Grand Solutions, Grant No. 6153-00010B). As well as by the Department of Clinical Medicine at Aarhus University.

## Declaration of interest

No authors have any conflicts of interest to declare.

Supplementary.

## Notes

### Competing Interest Statement

The authors have declared no competing interest.

### Funding Statement

This study was funded by by the Innovation Fund Denmark with a grant for the REMAP project (Grand Solutions, Grant No. 6153-00010B). As well as by the Department of Clinical Medicine at Aarhus University.

### Author Declarations

The scientific ethic committee of Region Midtjylland, Denmark gave ethicak approval for this work. (ID number: 65059)

